# Language Abnormalities in Alzheimer’s Disease Arise from Reduced Informativeness: A Cross-Linguistic Study in English and Persian

**DOI:** 10.1101/2024.03.19.24304407

**Authors:** Sabereh Bayat, Mahya Santai, Mehrdad Mohammad Panahi, Amirhossein Khodadadi, Mahdieh Ghassimi, Sahar Rezaei, Sara Besharat, Zahra Mahboubi, Mostafa Almasi, Morteza Sanei Taheri, Bradford C Dickerson, Neguine Rezaii

## Abstract

**INTRODUCTION:** This research investigates the psycholinguistic origins of language impairments in Alzheimer’s Disease (AD), questioning if these impairments result from language-specific structural disruptions or from a universal deficit in generating meaningful content.

**METHODS:** Cross-linguistic analysis was conducted on language samples from 184 English and 52 Persian speakers, comprising both AD patients and healthy controls, to extract various language features. Furthermore, we introduced a machine learning-based metric, Language Informativeness Index (LII), to quantify informativeness.

**RESULTS:** Indicators of AD in English were found to be highly predictive of AD in Persian, with a 92.3% classification accuracy. Additionally, we found robust correlations between the typical linguistic abnormalities of AD and language emptiness (low LII) across both languages.

**DISCUSSION:** Findings suggest AD linguistics impairments are attributed to a core universal difficulty in generating informative messages. Our approach underscores the importance of incorporating biocultural diversity into research, fostering the development of inclusive diagnostic tools.

## Background

The analysis of language production has emerged as a potent method for detecting cognitive abnormalities in Alzheimer’s disease (AD).^1–4^ The sensitivity of language in detecting AD extends to several years before the official diagnosis of the disease.^5,6^ Analysis of short language samples from a picture description task in cognitively unimpaired individuals has a higher accuracy of predicting incident AD compared to models based on neuropsychological test scores, demographic variables, and APOE results.^5^ Typical language abnormalities indicative of AD in English include using a higher rate of pronouns, shorter sentences, and an increased rate of adverbs.^2,4,7–10^ Despite its evident clinical value, the mechanism through which cognitive impairments affect language production remains poorly understood. Questions remain about whether the observed language changes are isolated distinct findings or if they are manifestations of a single core cognitive deficit. To address these questions, it is essential to determine the stage in language production where AD-related cognitive impairments emerge to result in characteristic language abnormalities.

There are two possibilities for how the cognitive impairments of AD can lead to the observed language abnormalities. The first is that these language abnormalities reflect impairments in cognitive capacities required to establish the surface structures of a language, such as the particular word order, assignment of grammatical gender, or other processes specific to a given language.^11,12^ For example, the reduced use of a specific word type in patients with AD (pwAD) might relate to the particular order in which that word type appears in a language, rendering it more vulnerable to being dropped. In this scenario, a different language with distinct structural rules may not exhibit similar impairment in the use of that word type. Alternatively, language abnormalities of AD may emerge from a deeper layer of language production where meaning is constructed before language-specific rules are applied. In this context, the language abnormalities of AD would not be bound to a particular language as they relate to a more universal aspect of language production: the formation of an informative message.

To attempt to answer these questions, we performed two stages of analysis. First, we investigated the extent to which language features of AD in one language can be transferred to another language with different surface structures. Specifically, we tested our first hypothesis that language features associated with AD in English would be able to reliably classify pwAD versus controls in Persian. Persian (also known as Farsi) is a typologically distant language relative to English, stemming from the Indo-Iranian branch of the Indo-European language family^13^ (Figure 1). Unlike English, which follows a subject-verb-object word order (SVO), Persian has the linear word order of subject-object-verb (SOV).^14–16^ One immediate outcome of this structure is that there are usually more words intervening between the subject-verb relationship in Persian compared to English (see Figure 2 for a comparison). In Persian, as in many SOV languages, adjectives are placed after the nouns they modify.^17^ Additionally, Persian pronouns have no grammatical gender.^18^ Persian also features a large, open-ended set of complex predicates comprising a non-verb element (e.g., a noun or adjective) followed by a light verb.^19,20^ Given the substantial structural disparities with English, Persian presents a compelling candidate language to pursue the objectives of this study. We extracted various language features indicative of AD in both English and Persian using transcriptions from participants as they described a picture. We then tested if the features indicative of AD in English are accurately able to classify pwAD who speak Persian. Poor generalizability might indicate that language abnormalities of AD stem from the inability to maintain surface features specific to English. Conversely, a high degree of transferability to Persian would suggest that the linguistic abnormalities of AD reflect disruptions at a deeper level of language production shared by both languages.

**Figure 1.**
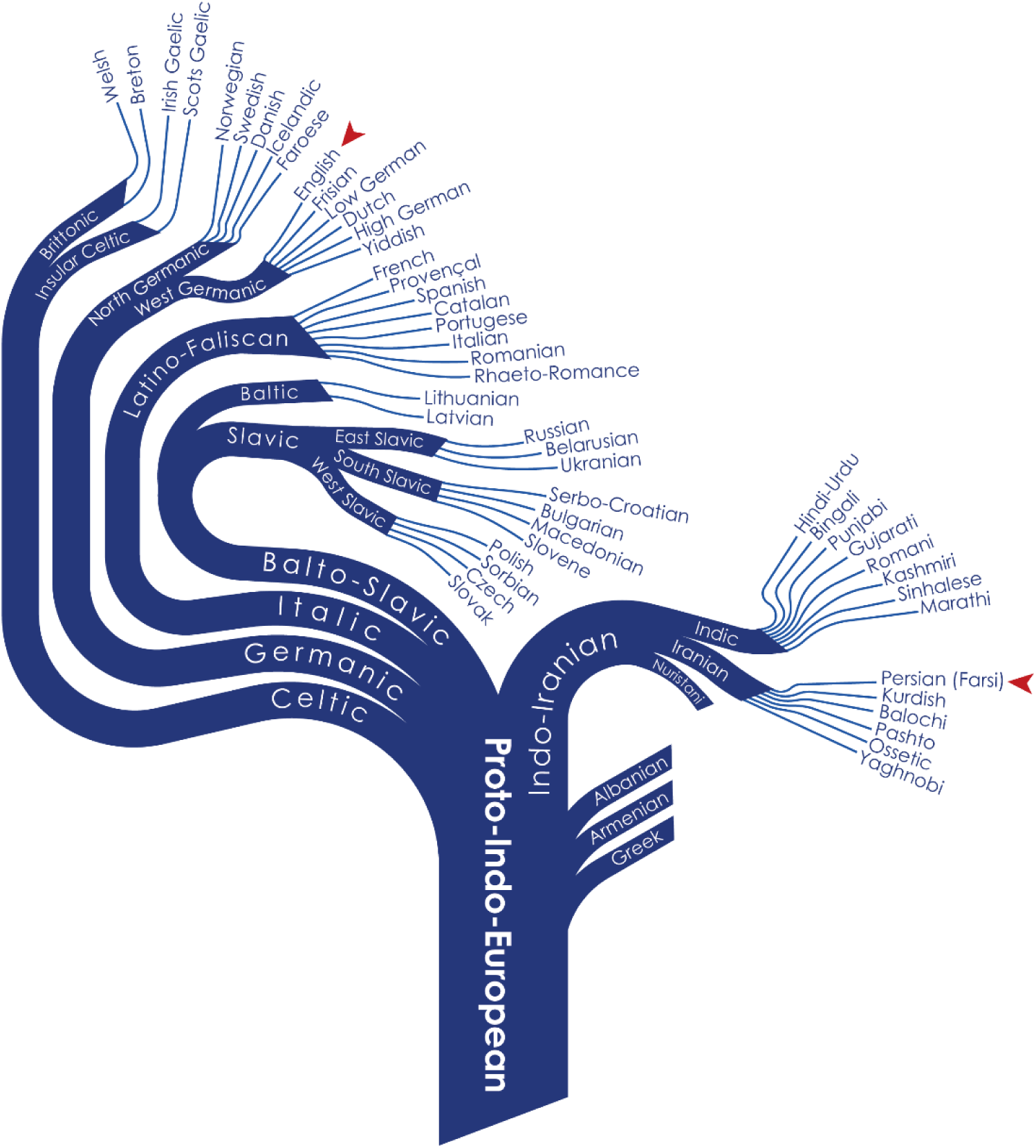
Persian (Farsi) belongs to a distant typological branch relative to English and originates from the Indo-Iranian branch of the Indo-European language family.

**Figure 2.**
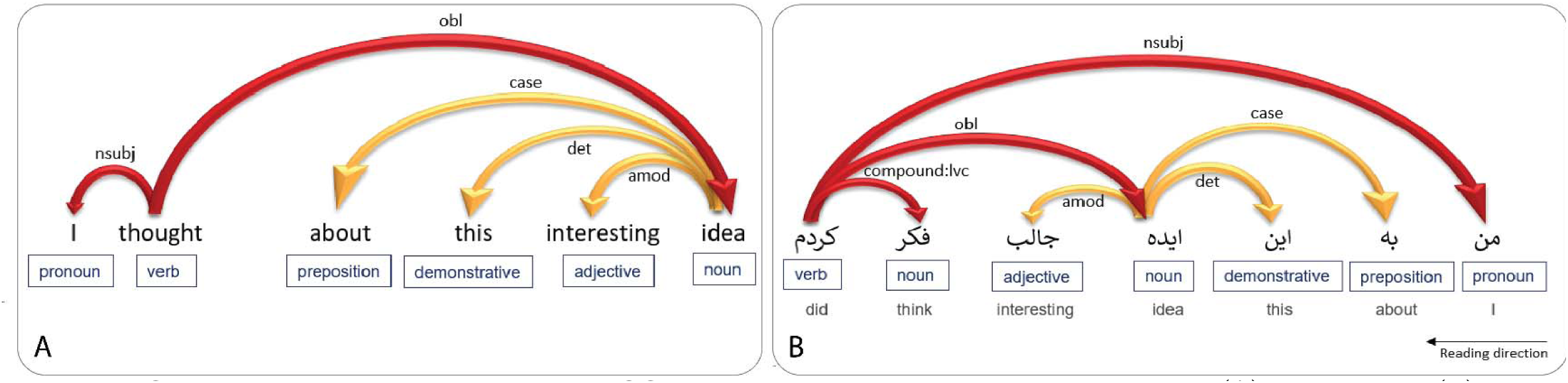
Syntactic parsing for determining POS tags and dependency relations in English (A) and Persian (B) of a similar sentence. The specific word order of each language results in varying distances between certain syntactic relations such as subject-verb relationship (nsubj).

Second, we hypothesize that the deeper level from which language abnormalities of AD originate is the stage of constructing an informative message. To test this hypothesis, we measured the informativeness of language produced by speakers of both languages and evaluated how informativeness correlates with language abnormalities of AD. To measure language informativeness, we used a novel metric we refer to as the Language Informativeness Index (LII) (see Box 1). Using a Large Language Model (LLM), LII measures the similarity of a target language sample to a highly informative description of the picture used in the language production task. We evaluated whether the typical language abnormalities of AD correlate with the LII measure, spanning both English and Persian. Strong correlations in both languages would provide evidence for the hypothesis that deficits in message formulation represent the pivotal stage at which language abnormalities in AD take root.

### Box 1. Language Informativeness Index (LII) and a review of measuring language emptiness in AD

A common method of measuring language informativeness, especially in language samples derived from picture description tasks, involves measuring the density of ideas. The literature varies in defining what constitutes an “idea.” Within this method, one approach is to evaluate the presence of specific items relative to language topics, referred to as content units or information units.^6,21–24^ For instance, when describing the Cookie Theft picture, key content units might include “mother” and “stealing cookies.” In a classic study, Croisile et al. identified 23 information units in the picture.^22^ This approach involves categorical scoring, assigning either a one or zero, without considering the varying salience of each information unit, such as “stealing cookies” versus “curtains”. Additionally, manual implementation of this method can be highly time-consuming. While automation is possible through word detection from a predefined list,^25^ the method remains sensitive to specific word choices. There are numerous ways to express a concept, particularly action items.

For instance, “stealing cookies,” “snatching sweets,” and “taking cookies without permission” all convey a similar idea, making it challenging to capture all variations using either manual or automated measures.

An alternative method for quantifying idea density involves counting propositions, typically defined as verbs, adjectives, adverbs, or prepositional phrases within every ten words of a text.^26,27^ In this approach, parts of speech that make insignificant contributions to the overall meaning still result in an increased informativeness score, as seen with adverbs in this sentence, “The kitchen looks so very messy”.

Other approaches involve measuring word frequency and lexical diversity.^28^ While these methods gauge the proficiency of patients in producing unique and informative words, they are not sensitive to the specific topic of speech. For example, a participant who employs a sophisticated and varied lexicon while recounting an old memory rather than addressing the required task of describing the Cookie Theft Picture might still receive a high informativeness score.

In this work, we introduce LII to measure language informativeness by measuring the similarity of participants’ language samples to a highly informative reference, here a detailed description of the cookie theft picture. We employed an artificial intelligence-based image-to-text tool to generate a reference text ensuring that it contains at least the 23 classic information units. Subsequently, we used a Large Language Model (LLM) to measure the similarity between each sample and the reference text (see Methods for more details). This metric allows for a graded scaling of informativeness rather than categorical scoring. Furthermore, LII is sensitive to the specific topic of language without being bound to particular word choices.

## Methods

### Participants

The English cohort. We obtained English samples from DementiaBank, a component of the TalkBank project.^29^ The dataset was collected at the University of Pittsburgh as part of the Alzheimer Research Program between 1983 and 1988 during a 5-year-follow up, with comprehensive information available in Becker et al. 1994.^30^ Inclusion criteria consisted of being older than 44 years old, having at least seven years of education, not having previous neurologic disorders, not taking neuroleptic drugs, having at least a score of 10 in Mini-Mental State Exam (MMSE) and being able to give informed consent. Participants were assessed using comprehensive examinations, including neuropsychological batteries, laboratory data, CT scans, and EEGs. From this dataset, we included 98 patients with probable AD and 86 age-matched healthy individuals. Since many participants had multiple language samples through the longitudinal approach, we only included their first sample. We selected the subset of pwAD from the DementiaBank in such a way that they match the MMSE and age of pwAD in the Persian cohort to maximize consistency across the two cohorts (Table 1).

**Table 1.** The demographic and clinical characteristics of pwAD and healthy individuals across the English and Persian cohorts.

|  | AD | Healthy individuals |
| --- | --- | --- |
| **English cohort** |  |  |
| **MMSE (mean ± SD)** | 18.4 (4.6) | 29.1 (1.1) p |
| **Age (mean ± SD)** | 67.9 (6.4) | 66.1 (6.3) |
| **Handedness (percent right-handed)** | - | - |
| **Sex (percent female)** | 66% | 58% |
| **Persian cohort** |  |  |
| **MMSE (mean ± SD)** | 17.5 (4.4) | 28.2 (1.3) |
| **Age (mean ± SD)** | 76.0 (5.6) | 68.5 (9.6) |
| **Handedness (percent right-handed)** | 100% | 96% |
| **Sex (percent female)** | 72% | 50% |

The Persian cohort. Twenty-five Persian-speaking pwAD and 27 age-matched healthy individuals were recruited from the Brain and Cognition Clinic in Tehran, Iran. A complete clinical history was taken from patients and their caregivers. Demographic features, clinical presentations, medical, and family histories were included in the interviews. A complete clinical examination was performed, emphasizing the assessment of motor features such as parkinsonism, praxis, language and speech, gait, and balance. The cognitive examination included Addenbrook’s Cognitive Examination-Persian version (ACE)^31^ and Mini-Mental State Exam (MMSE).^32^ Blood tests and the brain MRI of all patients were reviewed to confirm the diagnosis and rule out other medical conditions. The blood tests included complete blood count, biochemistry, renal and liver function tests, vitamin B12, 25-OH-D3, thyroid function test, syphilis, and HIV serologic test. Mild or Major Neurocognitive Disorders and Alzheimer’s disease were diagnosed by DSM-5 and NINCDS-ADRDA criteria^33^, respectively. In addition to obtaining a clinical history for all healthy individuals, the neurotypicality was confirmed using MMSE, brain MRI, or both. The research section of the Persian cohort was approved by the ethics committee of Iran University of Medical Sciences, which governs human subjects research in accordance with their guidelines. All participants provided written informed consent to participate in this study. We certify that the study was performed in accordance with the ethical standards as laid down in the 1964 Declaration of Helsinki and its later amendments. This study particularly champions biocultural diversity through the inclusion of participants from two linguistically and culturally distinct cohorts, thereby advancing the principle of avoiding ethnocentric biases in human data collection.

Language samples. Connected speech samples were obtained for both languages by asking participants to describe the Cookie Theft picture, a component of the Boston Diagnostic Aphasia Examination.^34^ Since the study concerns the lexicosemantic and syntactic features of language, disfluencies and false starts were removed based on a protocol previously described.^35^

Language features. We used Stanza, an open-source Python natural language processing toolkit that supports 66 human languages, including English and Persian, to extract part-of-speech (POS) tags and dependency relationships.^36^ Figure 2 shows syntactic parsing for determining POS tags and dependency relations in English and Persian. POS tags and dependency relations were normalized by dividing their raw counts by the total number of words each participant produced. Since this method results in hundreds of features, some of which are rarely used, we included those features used by at least 10% of all participants. We have previously provided a list of definitions and examples for features extracted from Stanza.^37^ In addition to the Stanza-derived feature set, we included word length, sentence length, average log frequency of all words and content words, total number of sentences, total number of words, and syntax frequency.^35^

Statistical analysis. To determine the correlation between language features and AD status (0 or 1), we used point bi-serial correlation. For classification, we used a binary logistic regression model. We employed a leave-one-out cross-validation (LOOCV) approach on our dataset to validate the model’s performance. In each iteration of the LOOCV, a single observation is set aside as the test data, and the remaining observations are used to train the model. To minimize the number of variables used in the logistic regression, we used Recursive Feature Elimination (RFE).^38^ RFE is a feature selection technique used in machine learning, particularly with logistic regression. It is a backward method of selecting predictors that starts with all available features, trains a model, evaluates feature importance, removes the least important features iteratively, and repeats this process until a desired number of features or a stopping criterion is met. RFE reduces dataset dimensionality, enhances model interpretability, and potentially improves generalization by focusing on informative features.

Language Informativeness Index (LII). LII measures the semantic similarity between a target text (produced by participants) and a reference text consisting of a meticulously crafted and highly informative description of the picture used for language production. We used a transformer-based model, the ‘bert-base-nli-mean-tokens’, from the Sentence Transformers library.^39^ This model is a specialized version of the BERT (Bidirectional Encoder Representations from Transformers) model, tailored for sentence-level embeddings and optimized for natural language processing tasks.^40^ The ‘bert-base’ variant is a more compact and efficient version of the model, featuring 12 transformer blocks, 768 hidden units, and 12 attention heads. We defined a function that takes two input texts to measure their similarity. Using the model, each text is first transformed into a high-dimensional vector representation (embedding). We then computed the cosine similarity between these embeddings, resulting in a similarity score ranging from 1 (identical in meaning) to –1 (completely dissimilar).

Reference text. Multiple approaches can be taken in generating the reference text, such as synthesizing a text using information units extracted from normative data^41^ or a predefined list of information units. Here, we used artificial intelligence-based image-to-text tools from OpenAI,^42^ ensuring that it contains all 23 standard information units.^22^ The list consists of 23 information units in four key categories: subjects, places, objects, and actions. The three subjects were the boy, the girl, and the woman. The two places were the kitchen and the exterior seen through the window. The eleven objects included cookie, jar, stool, sink, plate, dishcloth, water, window, cupboard, dishes, and curtains. Finally, the seven actions or facts were boy taking or stealing, boy or stool falling, woman drying or washing dishes/plate, water overflowing or spilling, action performed by the girl, woman unconcerned by the overflowing, and woman indifferent to the children. Below is the reference text in English.

“In the kitchen, a boy is standing on a stool. He is trying to steal cookies from a cookie jar on a cupboard shelf. The stool is tilted, so the boy is about to fall. A little girl is standing on the floor reaching up for some cookies. A woman, likely their mother, is washing or drying a plate with a dishcloth. The water is overflowing from the sink. The woman seems unaware or indifferent to the spilling water and the children’s actions. Outside the window, there is a little yard with a driveway. The window has curtains. There are some dishes on the counter.”

## Results

### The language features of AD in one language are generalizable to another language

We first determined the language features with the highest correlation with AD using point-biserial correlations for each language (Figure 3). In English, the language features with the highest correlation with AD include higher proportion of adverbs (r = 0.36, *P* < .001), shorter content words (r= –0.36, *P* < .001), higher proportion of pronouns (r = 0.35, *P* < .001), higher proportion of objects (r = 0.27, *P* < .001), higher content word frequency (log) (r = 0.26, *P* < .001), and reduced use of prepositions (r = 0.24, *P* < .001).

**Figure 3.**
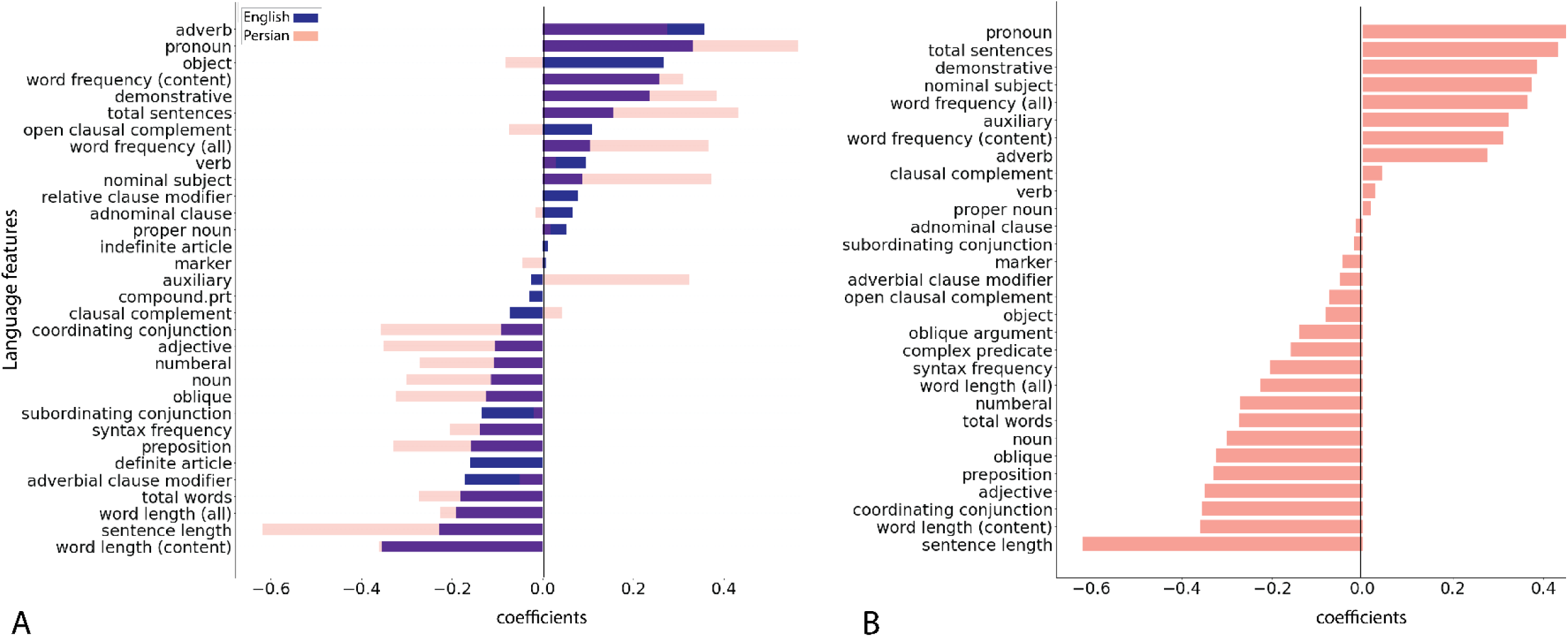
The correlation coefficients of language features with an AD diagnosis. A) The graph sorts the features based on the strength of correlation in English, with Persian features superimposed for direct comparison. B) The graph sorts the features based on the strength of correlation in the Persian cohort. Redundant features, such as adjectives as parts of speech (POS) and dependency relations, are shown only once to avoid repetition.

To test cross-linguistic transferability, we selected all features with a significant correlation with AD in the English cohort at the alpha level of 0.05 to build a classifier based on a binary logistic regression in the Persian cohort. We did not apply a correction for multiple comparisons at this stage since the RFE method inherently addresses the issue of multiple comparisons. RFE operates iteratively, evaluating the contribution of each feature to the classification model’s performance and eliminating the least significant features. By systematically ranking and removing features, RFE effectively prioritizes those with the greatest discriminatory power, reducing the potential for false discoveries. Therefore, the application of RFE minimizes the impact of multiple comparisons by selecting a parsimonious set of features that collectively optimize classification accuracy.

We applied LOOCV for cross-validation, which accounts for potential overfitting and provides a robust evaluation of our classifier’s generalization ability. Using the indicators of AD in the English cohort, LOOCV resulted in an average accuracy of 92.3% in classifying AD in the Persian cohort (average precision = 89% and average recall = 96%). The final feature set for AD classification after RFE included adverbs (as a POS), word length (content words), pronouns (as a POS), adverbial dependency relationship, content word frequency (log), demonstratives, sentence length, total words, and numerals (as a POS).

In a monolinguistic approach where we built a classifier based on the Persian feature set, rather than transferring from English, we reached an accuracy of 98.1% in classifying AD after applying RFE and LOOCV.

### LII shows that the language features of AD associate with reduced informativeness

First, we assessed the validity of LII by sequentially dropping individual information units from the reference text and evaluating the impact on LII. We manually eliminated each information unit from the text and compared its similarity to the original reference text. As expected, LII progressively declined as a function of sequentially eliminating information units, confirming the sensitivity of the index in gauging language informativeness (Figure 4).

**Figure 4.**
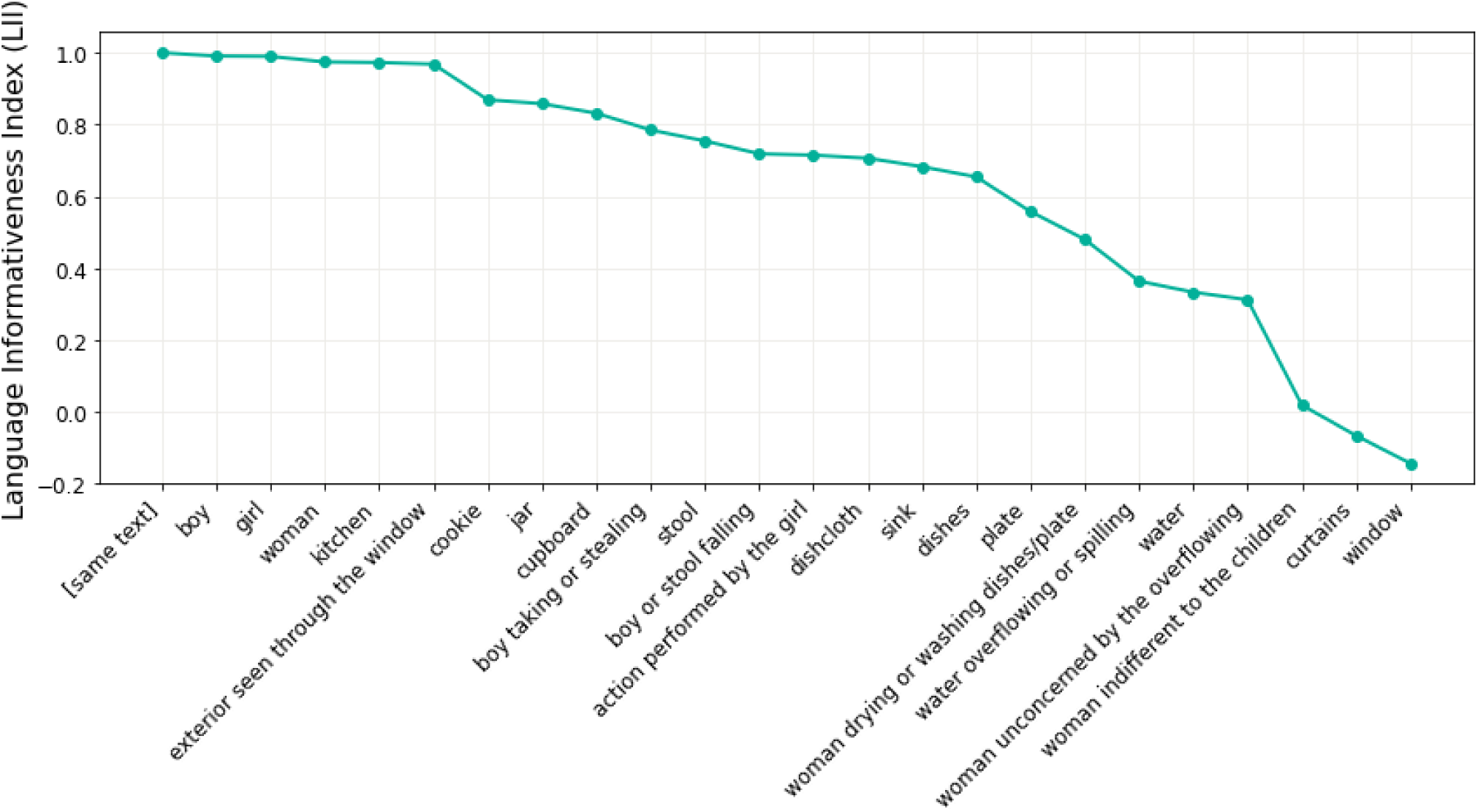
The reduction of LII as a function of excluding various content units in the Cookie Theft Picture. As each information unit is sequentially eliminated from the target text, its similarity to the reference text declines.

In the English cohort, pwAD produced text with a lower LII (mean = 0.84, SD = 0.12) compared to healthy individuals (mean = 0.90, SD = 0.05) t(127.7) = 4.61, *P* < .001. Similarly, in the Persian cohort, pwAD produced texts with a lower LII (mean = 0.98, SD = 0.005) than healthy individuals (mean = 0.99, SD = 0.005) t(49.65) = 4.03, *P* < .001 (Figure 5).

**Figure 5.**
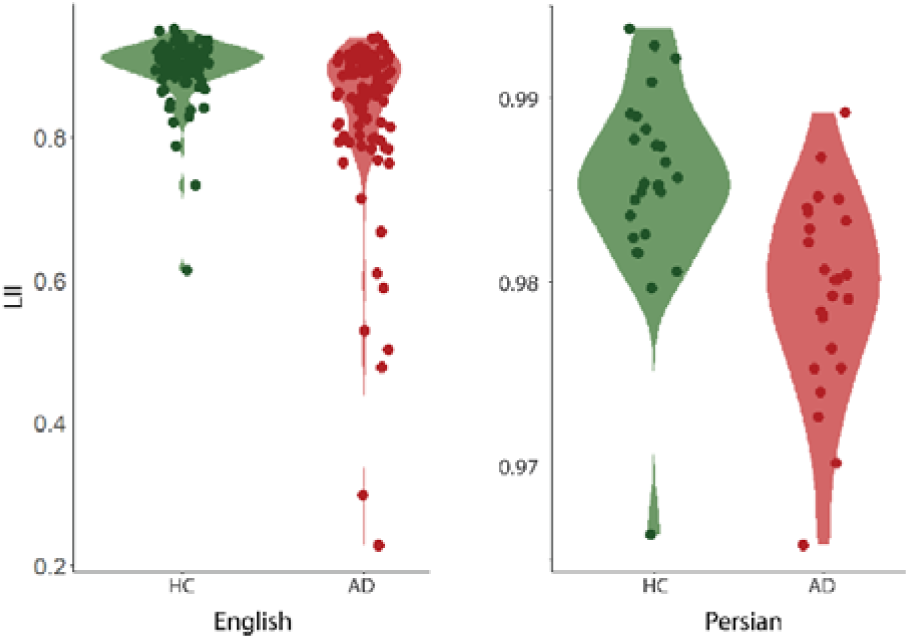
Violin plots of LII in pwAD and healthy individuals (HC) in English and Persian.

We then investigated the relationship between the typical language abnormalities of AD and LII. For instance, we sought to determine whether a high rate of the use of pronouns—which is a typical feature of AD—also correlates with low informativeness, suggesting the inability to produce informative messages as the origin of this language abnormality. We examined the amount of overlap between the pattern of correlations of language features with AD and the pattern of correlation of the same language features with low LII. As shown in Figure 6, we found a substantial overlap in the patterns of correlation of language features with AD and low informativeness. The ways in which the probability of AD changed with respect to language features are comparable with the ways in which low informativeness changes with respect to these language features. Where the AD graph shows an increase, the low informativeness graph follows in tandem, and this synchronized pattern persists in instances of decline. For instance, in English, the length of content words, rate of using adverbs, rate of using pronouns, and sentence length that had the highest correlation with AD (r = –0.36, *P* = .001, r = 0.36, *P* = .001, r = 0.33, *P* < .001, and r = –0.23, *P* =.002, respectively) showed comparable correlations with language emptiness (r = –0.33, *P* < .001, r = 0.30, *P* < .001 and r = 0.31, *P* < .001, and r = –0.25, *P* < .001, respectively).

**Figure 6.**
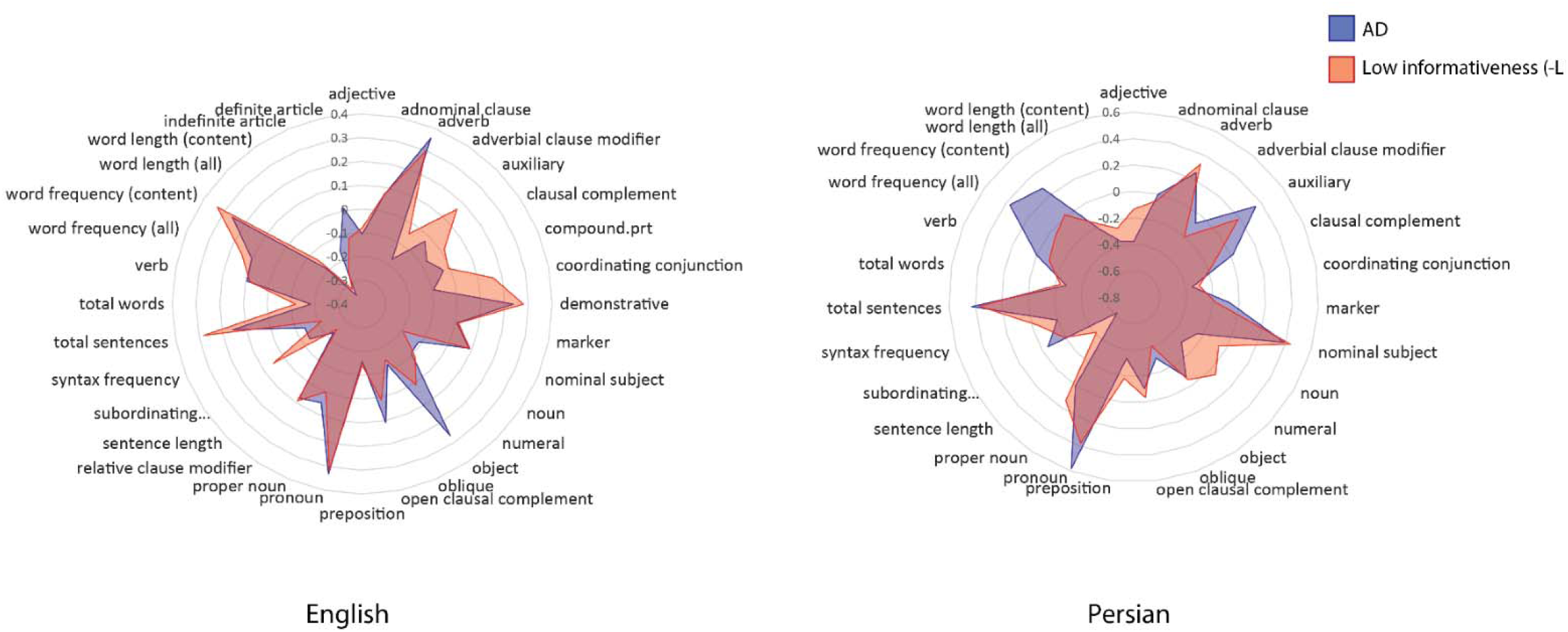
Radar charts depicting correlation coefficients between Alzheimer’s Disease (AD) and low informativeness (emptiness) across various linguistic features in English and Persian. Substantial overlap exists between the two variables with respect to linguistic features in both languages

Similarly, in the Persian cohort, sentence length, rate of using pronouns, and total number of sentences had the highest correlation with AD (r = –0.62, *P* < .001, r = 0.56, *P* < .001, and r = 0.43, *P* = .001, and r = –0.36, *P* = .009, respectively) and also showed comparable correlations with language emptiness (r = –0.43, *P* = .002, r = 0.37, *P* = .007, and r = 0.41, *P* = .002, respectively).

Interestingly, the rate of using objects, which showed poor transferability in predicting AD from English to Persian (Figure 3), had a correlation of almost zero with low informativeness (r = –0.01, *P* = .90). This finding suggests that features that are similar in English and Persian in predicting AD have a high correlation with emptiness and those with poor transferability have a minimal correlation with emptiness.

## Discussion

In this work, we sought to elucidate the psycholinguistic foundations of language impairments in the connected speech of people with Alzheimer’s disease. Although numerous studies have underscored the diagnostic utility of language analysis in identifying people with or who will develop AD, the mechanisms underlying these language abnormalities remain poorly understood. Language production is a complex process that begins with generating a thought-level message. Once the message is ready for expression, the formulation process unfolds, which entails accessing lexical elements, specifying grammatical relations, and mapping the resultant output onto inflectional and phrasal structures.^11^ It is currently unclear how the hallmark cognitive impairments of AD affect the process of language production.

To delineate this mechanism, we contrasted two possibilities. The first possibility is that the linguistic abnormalities of AD could be attributed to a deterioration in cognitive functions necessary to establish a given language’s specific morphosyntactic rules. The second possibility is that the language anomalies might indicate a more fundamental and universal disruption in the language production process at the level of message formation, transcending surface structures specific to a given language. By comparing native speakers of English and Persian, which are languages with considerable structural differences, we demonstrated that the English linguistic indicators of AD could be used to construct a classification model for AD in Persian with an accuracy of 92.3%, supporting our first hypothesis. The high degree of transferability of language indicators of AD from English to Persian suggests that these indicators likely do not stem from a breakdown in language-specific morphosyntactic rules. This conclusion is reinforced by observing shared linguistic indicators of AD across English and Persian, such as reduced use of adjectives despite their different syntactic ordering.

Second, we hypothesized that the primary deficit in AD-related language abnormalities is an inability to construct informative messages, which we tested using the Language Informativeness Index (LII), a novel metric for informativeness. We found robust correlations between typical language indicators of AD and language emptiness in both English and Persian. Crucially, there was substantial overlap in how informativeness and likelihood of AD changed with respect to language features. The few language features that did not show a comparable relationship with LII and AD, such as the rate of using objects, had poor transferability across languages in classifying AD. Our findings suggest that the inability to form a clear, informative message might be the origin of the typical language features of AD, such as increased use of pronouns, increased use of adverbs, shorter words, shorter sentences, increased use of demonstratives (e.g., “this” or “that”), high word frequency, and decreased usage of numerals.

Although language emptiness has been a widely acknowledged finding in AD literature,^21,26,43,44^ several critical research gaps hindered the emergence of a comprehensive understanding of the neurolinguistic underpinnings of language impairments in AD. A major limitation has been the absence of a rigorous method for quantifying language emptiness, which in turn hindered investigations to establish a clear relationship between diminished informativeness and the language indicators of AD. Consequently, the language abnormalities observed in AD have been viewed as separate findings rather than a reflection of a fundamental problem with the generation of informative messages. The informativeness of language, arguably its most fundamental element, is challenging to quantify. Effective measurement needs to consider both the richness and variability of words, as well as their relevance to the topic of speech. Previous approaches often focused on some but not all of these essential components. In this work, we introduced LII as a fully automated approach that provides a graded scoring of language informativeness. The metric is sensitive to the richness, variability, and relevance of words without being bound to a particular wording.

Another notable hurdle in understanding the mechanism of language abnormalities of AD is the scarcity of cross-linguistic approaches in the field. A predominant focus on a single language fails to distinguish between various alternative explanations, such as whether abnormalities stem from surface structures of the language or deeper layers of message formation. These ethnocentric approaches can be limited in their scope to provide new insights^45^ and might even result in misconceptions about language phenomena (for example, see references^46,47^ for discussions about perspectives on nonfluent aphasia that may stem, at least in part, from an ethnocentric approach to language analysis).

When cross-linguistic similarities are observed in comparative linguistics studies, several potential explanations are considered.^13^ The explanations include pure chance, borrowing of linguistic elements due to language contact, close branching in a language family tree, or linguistic universals—features common across languages due to inherent properties of human language capacity. The high transferability rate of 92% from English to Persian in classifying AD makes pure chance an unlikely explanation. Additionally, by selecting two linguistically distant languages, we minimized the influence of cultural and typological similarities.

Therefore, we propose that the observed commonalities in AD language indicators between English and Persian stem from a shared underlying feature: poor message formation. Language universals are thought to have biological foundations stemming from a genetically determined language faculty shared among speakers of all languages.^12^ Additionally, as speakers of different languages share a common cognitive apparatus, they are expected to possess a set of foundational properties that underlie language production and comprehension.^48^ Here, we suggest that message generation is one such biologically plausible language universal that is affected in AD, resulting in its typical language features across English and Persian (and potentially other languages).

In conclusion, our study offers a comprehensive understanding of the psycholinguistic underpinnings of language abnormalities in Alzheimer’s disease. By employing a robust metric of language informativeness based on artificial intelligence and embracing a cross-linguistic approach, the work establishes links between the typical language abnormalities of AD and the inability to produce informative messages. This connection highlights a universal aspect of language production as the affected stage of language production in AD, transcending specific linguistic structures. Future work is needed to expand the approach to other languages, particularly those belonging to entirely different family trees. In addition to enhancing our understanding of the psycholinguistic deficits of AD, this approach could lead to the development of diagnostic tools across various languages, ultimately fostering health equity and a deeper appreciation of biocultural diversity.

## Data Availability

All English samples are available through Dementia Bank which is publicly accessible. The Persian samples can be accessed through a reasonable request to the senior author of the manuscript at.

## Acknowledgment

We thank the support from Harvard Catalyst, The Harvard Clinical and Translational Science Center (National Center for Research Resources and the National Center for Advancing Translational Sciences, National Institutes of Health Award UL1 TR002541) for its biostatistician consultation service. We also thank Mahan Rezaei for transcriptional services in Persian. This work was supported by Alzheimer’s Association Clinician Scientist Fellowship (AACSF) 2022A015154 and MGH Screening Technologies in Primary Care Innovation Fund (PCIF) 2023A063002, as well as National Institutes of Health grants R21 DC019567, R21 AG073744, and R01 NS131395.

